# The Menstrual Practice Needs Scale Short Form (MPNS-SF) and Rapid (MPNS-R): Development in Khulna, Bangladesh, and validation in cross-sectional surveys from Bangladesh and Uganda

**DOI:** 10.1101/2024.01.22.24301625

**Authors:** Julie Hennegan, Md. Tanvir Hasan, Tasfiyah Jalil, Erin C Hunter, Alexandra Head, Abdul Jabbar, Arifa Bente Mohosin, Nigar Sultana Zoha, Muhammad Khairul Alam, Laura Dunstan, Sabina Akter, Afreen Zaman, Adrita Kaiser, Calum Smith, Lillian Bagala, Peter S Azzopardi

## Abstract

**Objectives:** Develop and validate short and rapid forms of the 36-item Menstrual Practice Needs Scale (MPNS-36).

**Design:** Item reduction prioritised content validity and was informed by cognitive interviews with schoolgirls in Bangladesh, performance of scale items in past research, and stakeholder feedback. The original MPNS-36 was revalidated, and short and rapid forms tested in a cross-sectional survey. This was followed by further tests of dimensionality, internal consistency, and validity in multiple cross-sectional surveys.

**Setting and participants:** Short form (MPNS-SF) and rapid form (MPNS-R) measures were developed in a survey of 313 menstruating girls (mean age=13.51) in Khulna, Bangladesh. They were further tested in the baseline survey of the Adolescent Menstrual Experiences and Health Cohort, in Khulna, Bangladesh (891 menstruating girls, mean age=12.40); and the dataset from the MPNS-36 development in Soroti, Uganda (538 menstruating girls, mean age=14.49).

**Results:** The 18-item short form reflects the six original subscales, with the four core subscales demonstrating good fit in all three samples (Khulna pilot: RMSEA=0.064 90%CI 0.043-0.084, CFI=.94, TLI=.92. Cohort baseline: RMSEA=0.050 90%CI 0.039-0.062, CFI=.96, TLI=.95. Uganda: RMSEA=0.039 90%CI 0.028-0.050, CFI=.95, TLI=.94). The 9-item rapid form captures diverse needs. A two-factor structure was the most appropriate but fell short of adequate fit (Khulna pilot: RMSEA=0.092 90%CI 0.000-0.158, CFI=.93, TLI=.89). Hypothesised associations between the MPNS scores and other constructs were comparable between the MPNS-36 and MPNS-SF in all populations, and replicated, with attenuation, in the MPNS-R. Internal consistency remained acceptable.

**Conclusions:** The MPNS-SF offers a reliable and valid measure of adolescent girls’ menstrual hygiene experience while reducing participant burden, to support implementation and improve measurement in menstrual health research. The MPNS-R provides a brief measure with poorer structural validity, suited to including menstrual health within broader water, sanitation and hygiene or sexual and reproductive health research.

## Introduction

Quantification of adolescent girls’ menstrual health needs – essential to population monitoring and evaluating the effectiveness of interventions - has been limited by a lack of measures for core concepts.[1, 2] The Menstrual Practice Needs Scale (MPNS-36) [3] was published in 2020 to address this gap and has seen rapid uptake in research and practice.[4–12] However, this comprehensive scale is 36-items in length, presenting a barrier to implementation in short needs assessments or multi-component surveys. Stakeholders have requested a short form to enable greater uptake.

The MPNS-36 measures respondents’ experience of menstrual blood management during their last menstrual period.[3] It assesses the extent to which an individuals’ needs for menstrual absorbents, disposal, spaces for changing and laundering reusable materials were met. In doing so, it provides a participant-centred measure of a key requirement for menstrual health outlined in the definition: *“women, girls, and all other people who experience a menstrual cycle are able to care for their bodies during menstruation such that their preferences, hygiene, comfort, privacy and safety are supported.”*[13] As such, it is also a measure of menstrual hygiene experience.[14] Individual scale items can be used to understand the experiences and needs of respondent population (e.g., [9]), while changes in total and sub-scale scores can be used to evaluate the effectiveness of menstrual health interventions (e.g., [8, 11]). The scale can also be used in research to test associations between risk and protective exposures and menstrual experience, and to quantify relationships between unmet menstrual management needs and consequences for women’s and girls’ health, social and educational outcomes (e.g., [15]). This broad range of uses must be considered in developing a shorter form.

The MPNS-36 is comprised of 36 items which can be delivered as personalised statements for self-report or as questions for enumerator administered surveys. Each item asks about the frequency of experience during the last menstrual period on a four-point Likert scale from *never* to *always*. Adolescent sub-scales include material and home environment needs, material reliability concerns, change and disposal insecurity, and transport and school environment needs, along with reuse needs and reuse insecurities. The scale was developed to assess experiences across the breadth of blood management practices and perceptions of the environments used for menstrual management, with practice domains derived from a systematic review of qualitative studies of menstrual experiences in low-and-middle-income countries (LMICs).[16] This comprehensiveness contributed to the length of the measure, but also provides a granular picture of population needs. In evaluating interventions, the MPNS ensures that management tasks that may not be the target of the intervention are assessed. For example, many interventions focus on delivering menstrual products. Without considering individuals’ experiences of disposal for single-use products or laundering for reusables, evaluations are likely to provide an incomplete and inaccurate picture of the effect of interventions on menstrual experience.

### The present study

We aimed to provide short and rapid versions of the MPNS to meet the needs of different users, and to compare the performance tools at shorter lengths. For the short form we aimed to halve the length of the MPNS-36, and to halve this again for a rapid version.

Guiding principles for item reduction were set a-priori based on past research and theory. First, we prioritised content validity and retaining the breadth of experiences assessed through the measure above structural validity. Item selection was not driven by item factor-loadings alone. As noted above, the experiences measured draw on systematic review of qualitative research and ensure that intervention evaluations capture holistic menstrual management experiences. Second, single items from the MPNS have been included as part of recommended indicators for national and global monitoring of menstrual health and hygiene.[17, 18] We prioritised retaining these in shorter forms to enable comparability of data collected using the scale with national data. Third, we decided a-priori to retain items in the MPNS that capture experiences separately relating to the home and school environments. Research has consistently highlighted differing experiences at home and at school (for adolescents) or work (for adults [19]), and studies using MPNS data have consistently shown differing experiences of menstruation in these settings.[6, 9] While duplicate items contribute to length of the scale, they are useful for policy and practice. In needs assessment, they highlight areas of greatest need, while in evaluations differences in items over time or between study sites can provide feedback on environments improved by the intervention. Fourth, we prioritised retaining a balance of positively and negatively orientated items in shortened versions. Assessments of MPNS dimensionality have consistently found that items capturing positive appraisals of experience such as satisfaction with the available changing facilities or having enough menstrual products, load on separate but correlated factors to those capturing insecurities such as worries about privacy or leaking.[3, 4, 6] As noted in the original development, including both positively and negatively framed experiences balances framing in administering the items with participants and offers a more nuanced assessment of experience. Combined scores across positively and negatively scored items have demonstrated stronger relationships with hypothesised correlates than these item sets alone across multiple studies.[3, 6, 15]

## Methods

The development and assessment of the short form (MPNS-SF) and rapid (MPNS-R) was undertaken over multiple phases and drew on stakeholder feedback and past research outlined in the Background. First, we re-validated the full MPNS-36 in Bangladesh using cognitive interviews with girls to assess item comprehension and quantitatively through a pilot cross-sectional survey in Khulna, Bangladesh. Second, we developed a candidate short form. Items were prioritised drawing together findings from the same cognitive interviews with girls using participatory activities to understand their perspectives on item importance, factor loadings for items observed across existing studies, use of MPNS items as national indicators, and theoretical considerations outlined in the Background. Third, the factor structure of the candidate short and rapid forms were tested and validated using the Khulna pilot survey data. Fourth, MPNS-SF and MPNS-R validity and reliability were appraised in the original MPNS development dataset (Soroti, Uganda), and the baseline of the Adolescent Menstrual Experiences and Health Cohort (AMEHC) study[20] in Khulna, Bangladesh (Khulna Cohort, Bangladesh).

### Study samples and data collection

#### Khulna Pilot, Bangladesh

Prior to the launch of the AMEHC study, a preparatory mixed-methods research program was undertaken to understand the menstrual health challenges in the setting and to refine cohort measures. Khulna District, specifically Khulna City Corporation (urban) and Dumuria Upazilla (rural), were selected as sites for the AMEHC study through collaboration among the cohort research partners.[20] Study measures, including the MPNS-36, were translated from *English* to *Bangla* by bilingual research team members and reviewed in group sessions. Cognitive interviews.

Ten cognitive interviews focused on the MPNS were undertaken with 20 adolescent schoolgirls in pairs in September 2022, facilitated by a trained female interviewer and lasting 60-90 minutes. Girls participated from five of the six schools that had been purposively selected as part of the qualitative phase of research. Schools had a strong relationship with the study NGO partner and included co-educational, single-sex, and madrassa (religious education) school types. Participants were provided with a written copy of the MPNS questions, grouped by sub-scale, and discussed their answer, rationale and understanding of each item. Most groups engaged with half of the survey items (3 sub-scales) to avoid fatigue. At the end of each sub-scale, participants were provided with flashcards of each question and asked to sort them into three categories ‘most important’ ‘important’ and ‘least important’. The interviewer used the interview audio-recording to produce a written English summary of participant rationale for item responses and prioritisation. Daily debriefing sessions amongst the research team refined the *Bangla* translation of items, and updated translations were deployed in the following day’s interviews.

### Survey

A pilot survey was undertaken across 10 purposively selected schools with girls attending Classes 6-9 and aged 12-16 in October 2022. A target sample of 360 participants provided 10 participants per MPNS item. Female enumerators received 5 days of training and administered the survey verbally, entering responses into tablets and uploading to the BRAC James P Grant School of Public Health KoboToolbox server. Girls were provided with a written copy of the survey if they wished to follow along and read the questions for themselves. The printed survey included the visual response tool for the MPNS depicting the four response options.[3] Surveys lasted 45-60 minutes for post-menarche participants.

Cognitive interviews and the survey followed sensitization workshops at each school notifying teachers and parents about the study. Parents/Guardians provided written consent, and girls provided written assent to participate.

#### Khulna Cohort, Bangladesh

Methods and sampling for the AMEHC study are detailed elsewhere.[20] In brief, 101 schools from Khulna City Corporation and Dumuria Upazilla were selected using a proportional random sampling approach to achieve a representative sample of adolescent girls attending co-educational, single-sex and madrassa schools. All girls attending Class 6 were eligible to participate. Following six days of training, surveys were administered by female enumerators. Participants were provided a written copy of the survey. Data was collected between February and March 2023. Surveys with girls who had reached menarche were an average of 30 minutes duration. Parents/Guardians provided written consent, and girls provided written assent to participate.

#### Soroti, Uganda

Methods and results of the original MPNS-36 development and validation in Soroti, Uganda have been published previously.[3] The dataset is publicly available. Twelve schools engaged with the partnering NGO were recruited. Adolescent girls attending primary school class levels 4-7 were included, with most participants from primary class levels 5-6. Paper copies of the survey in English were provided to groups of no more than six girls. Trained female enumerators facilitated the survey, providing verbal translation of each item in Ateso, with participants indicating their own responses on the paper survey. Data collection was undertaken between March and May 2019. Surveys lasted 75-90 minutes. Schools consented to participation and informed parents through parent-teacher meetings with the option to opt-out of the study. Girls provided written assent.

## Measures

### Sample demographics and menstrual practices

Participants self-reported their age, class level and the materials they used as menstrual absorbent. In all three data collections, girls self-reported if they washed and reused any menstrual materials during their last period. In both Bangladesh data collections, girls also self-reported if they attended school during their last menstrual period, and if they changed their menstrual materials at school. These were used as subsequent eligibility criteria for MPNS items.

### The MPNS

The full MPNS-36 was administered to girls in the Khulna pilot. The subsequent cohort baseline used the short-form items only. Both Bangladesh surveys used the interview version of the MPNS, which presents each item in question format, for example *“During your last menstrual period, were your menstrual materials comfortable?”.* The full set of questions are displayed in tables in the results section. Participants provide responses on a 4-point Likert scale: never, some of the time, most of the time, always. These terms were determined to be appropriate in the *Bangla* translation, however training with enumerators also highlighted that the middle options could also be interpreted as ‘less than half of the time’ and ‘more than half of the time’. Prior to asking the MPNS items, in both the pilot and cohort surveys, enumerators administered a brief exercise to familiarise girls with the response options. This included asking girls about daily activities such as *“Over the past week, how often did you have street food?”*. Girls were also presented with a printed copy of the survey which displayed the response options and MPNS visual response tool [3]. Participants were asked to respond to MPNS items concerning the disposal (Items 12-15) of menstrual materials if they reported disposing of materials during their last period, including disposing of single-use or reusable products at the end of their life. They were asked to respond to questions about laundering materials if they reported washing and reusing any material during their last period (items 29-36). Girls who reported that the ‘never’ changed their materials at school during their last period were not asked questions about the experience of changing materials at school (items 25-28).

In Soroti, Uganda, as part of the original MPNS-36 development, girls completed a longer set of candidate scale items. These were delivered as statements which girls responded to on their own survey, for example *“During my last menstrual period, my menstrual materials were comfortable”* with response options: never, sometimes, often, always.

MPNS sub-scale and total score are calculated using the mean of included items. Positively worded items are scored from 0-3 and negatively worded items from 3-0. Higher MPNS scores represent more positive experiences of menstrual blood management.

#### Hypothesised correlates

Mental health was assessed across all three datasets using a translated version of the Depression Anxiety Stress Scale (DASS-21) [21, 22]. In the Khulna cohort baseline and study sample in Soroti, Uganda, only the Depression and Anxiety sub-scales were used. A continuous total score of included items was calculated. Higher scores represent greater endorsement of depression and anxiety items and thus poorer psychological health. Although more research is needed, the DASS has been widely used across contexts, and has exhibited a bifactor structure scoring across items.[23] A *Bangla* version of the DASS-21 has previously been validated among adults in Bangladesh.[24]

Confidence to manage menstruation was assessed using similar self-report items across all three datasets. Participants were asked to report their agreement on a 4-point Likert scale (very unconfident, unconfident, confident, very confident) to the question relevant to home and then to school: *How confident do you feel that you can manage your menstruation [pad yourself, change your materials, dispose of them or wash and dry them] when you are at home?* In Bangladesh, to remain consistent with past research,[19] confidence at home was dichotomised to compare girls who reported being ‘very confident’ to those giving other responses, whereas confidence at school was dichotomised to compare girls who reported being ‘very confident’ or ‘confident’ to those who were not confident. In the Soroti dataset, girls who reported feeling ‘confident’ at both home and school were compared to ‘not confident.’ Groupings were maintained for comparability with the original MPNS-36 development.

School participation was assessed through two self-report items in the Bangladesh studies, one capturing self-reported absence from school during the last menstrual period, and the second asking girls to report if they had trouble participating in school, such as participating in class, due to their last menstrual period. This question asked for a yes or no response, and was aligned with new recommendations for monitoring menstrual health and hygiene.[17, 18] Participation during menstruation was not included in the study in Soroti, so only self-reported absence during the menstrual period was used.

### Analyses

Quantitative analyses were undertaken in Stata 17 and in *R* version 4.3.1. Revalidation of the MPNS-36 drew on girls’ reflections in cognitive interviews used to refine translation and check interpretability of the questions. Quantitative revalidation was undertaken in the Khulna Pilot data. Descriptive statistics were used to summarise sample responses to each MPNS item and identify missing data. Confirmatory factor analysis (CFA) using the *lavaan*[25] package for R was undertaken using a diagonally weighted least squares estimator (DWLS). We considered RMSEA ≤0.08 as indicative of a fair fit, and Comparative Fix Index (CFI) and Trucker-Lewis Index (TLI) ≥0.90 as indicative of acceptable fit (with CFI and TLI ≥0.95 indicative of close fit).[26] Factor loadings ≥0.30 were considered acceptable. Scaled estimates are presented. Internal consistency was assessed using Cronbach’s alpha, and ordinal alpha calculated using the polychoric correlations given the four-point response scale.[27] Validity was assessed by exploring associations between the MPNS-36 total and sub-scales with constructs hypothesised to be related.

To develop the short and rapid forms, girls’ perception of item importance from cognitive interviews were integrated with a-priori defined priorities for item selection. Girls scoring of the importance of items in cognitive interviews were used qualitatively and in the context of interview discussion of these decisions.

The dimensionality, internal consistency and validity of the short form was tested in the Khulna Pilot survey following the same procedures used for the MPNS-36 revalidation described above. For tests of dimensionality (structural validity) we first undertook CFA, hypothesising the original sub-scales would be replicated in the short form. To supplement this assessment, we also undertook exploratory factor analyses (EFA) using the polychoric correlation matrix and oblique rotation (*promax*) to investigate alternative structures, however the original was the best fit for the data for the short form. To test the dimensionality of the rapid form, we undertook EFA, and CFA testing hypothesising one and two-factor structures.

Tests of dimensionality of the short and rapid forms using CFA were then replicated in the Khulna Cohort Baseline, Bangladesh and Soroti, Uganda datasets. We note that DWLS requires complete data and so analyses reflect the sample of participants with no missing items. In the dataset from Soroti, Uganda we used the first multiple imputation data generated using chained equations with the *mice* package[28] in R from the original study.[3] Internal consistency using Cronbach’s alpha and ordinal alpha, along with tests of associations with hypothesised correlates were undertaken. Correlation coefficients or binary logistic regressions were used as appropriate to the distribution of the outcome.

### Patient and public involvement

NGO practitioners and researchers as the potential users and audience for the measure informed original development of the MPNS. Feedback from users shared with the development team informed considerations for short and rapid form development and cognitive interviews with girls were undertaken to support item selection. Community consultation for the broader AMEHC study was undertaken (see [20]) but this did not include focus on the MPNS.

## Results

### Survey sample characteristics

Sample characteristics of the three quantitative datasets used in the study are presented in Table 1.

**Table 1.** Sample characteristics for included populations.

|  | **Khulna Pilot,**<br>**Bangladesh** | **Khulna Cohort,**<br>**Bangladesh** | **Soroti, Uganda** |
| --- | --- | --- | --- |
| No. menstruating girls | 313 | 891 | 538 |
| Age range | 12-16 | 10-16 | 11-19 |
| Age mean (SD) | 13.51 (SD=1.13) | 12.40 (SD=0.94) | 14.49 (SD=1.20) |
| Exclusively used disposable commercial pads during last menstrual period % (n) | 61.98 (194) | 47.19 (420) | 29.37 (158) |
| Used reusable materials during the last period % (n) | 22.36 (70) | 41.01 (365) | 54.49 (291) |
| Changed materials outside the home (most likely at school) during their last period | - | - | 87.71 (472) |
| Changed materials at school during the last menstrual period | 31.52 (87) | 14.78 (116) | - |

### Re-validation of the MPNS-36 in Bangladesh

Twenty girls across classes 6-9 and aged 13-16 participated in cognitive interviews testing comprehension of MPNS items, translation, and informing the subsequent short-form development. Initial cognitive interviews highlighted easily understood questions, and those that required further amendments to translation. Most translation improvements were grammatical, relating to the ordering of sentence content. There were also modifications based on individual words. For example, “comfortable” had multiple translations in *Bangla* dependent on physical or emotional comfort. MPN1 refers to physical comfort, whereas items such as MPN10 referred to “feeling comfortable’” in English and the selected translation prioritised mental safety/peace comfort. By the conclusion of early translation modifications, all MPNS items were well-understood by respondents. In interviews, participants described varied circumstances and preferences that influenced their response selection. Quotations are presented in Supplementary Materials 1.

The MPNS-36 exhibited strong performance in the survey data collected in the Khulna Pilot in Bangladesh. The original MPNS-36 factor structure was an acceptable fit for the data (CFI=0.924, TLI=0.927, RMSEA=0.075, 90%CI=0.060-0.090). Sub-scales and total scale exhibited good internal consistency (total scale α=0.86) and the total and sub-scales showed multiple expected relationships with other constructs, including mental health, participation in school, and confidence managing menstruation at school. Full tables and text reporting the revalidation are presented in Supplementary Materials 2. This supplement also reports the responses to each survey item in this population.

### Short and rapid form item selection

There was agreement across the cognitive interviews that the experiences captured in the MPNS items were important and relevant for girls. There was substantial inconsistency across the interviews in the items rated as most or less important. In discussions, girls described that items were likely to be more important for different individuals, depending on circumstances. For example, groups consistently rated safety at school (MPNS 28) as a less important item because they did not personally have concerns about their safety in school toilets. However, safety at home (MPNS21 and 22) were rated by multiple pairs as “very important”, with girls’ emphasising that if this was a concern for girls, it would be a top priority. Pairs who did not wash and reuse materials did not see the relevance of these items, while those with difficulties rated this as ‘very important’; particularly privacy for washing and drying absorbents. In the four sub-scales that apply to all participants, the most highly prioritised items were:

- Material and home environment needs: comfortable menstrual materials (MPNS1) and being able to wash hands (MPNS11). Hand washing was particularly relevant in the context where menstrual items are considered unclean, and participants described washing their hands after having any contact with items related to menstruation.
- Change and disposal insecurity: being worried others would see disposed menstrual materials (MPNS14) and privacy for changing menstrual materials at home (MPNS20).
- Material reliability concerns: being worried materials would leak (MPNS5).
- Transport and school environment needs: being able to change menstrual materials when desired at school (MPNS23).

The rationale underpinning item prioritisation and selection for the short and rapid forms are summarised in Table 2. Content validity was the highest priority when reducing the item set. While more similar items, related to more similar practices, often exhibited higher factor loadings and enhanced model fit, we prioritised selecting a single item that represented each practice experience and removed items that provided different perspectives on a similar aspect of blood management experience. Items were also prioritised based on (a) their selection as indicators for national and global monitoring, (b) item performance in past data collections, for example while items 8 and 9 capture unique experiences of transporting materials they have cross-loaded and created problems for scale factor structure,[6] and (c) girls’ perspectives on item importance in cognitive interviews.

**Table 2.** Rationale for prioritised and selected short form (SF) and rapid (R) items.

|  | Item | Priority | SF | R | Rationale |
| --- | --- | --- | --- | --- | --- |
| 1 | Were your menstrual materials comfortable? | High | X |  | Priority for assessing material quality/suitability. Girls rated highly in cognitive interviews. |
| 2 | Did you have enough of your menstrual materials to change them as often as you wanted to? | Essential | X | X | Included in shortlist of indicators for national and global monitoring of menstrual health. |
| 3 | Were you satisfied with the cleanliness of your menstrual materials? | Lower |  |  | Greater potential for social desirability bias if participants are embarrassed to report dissatisfaction with cleanliness. Lower ratings in cognitive interviews. Often dictated (as described in cognitive interviews) by being able to change disposables regularly or ability to clean reusables, which are captured in other scale items. |
| 4 | Could you get more of your menstrual materials when you needed to? | Lower |  |  | Both MPNS4 and MPNS7 capture access to materials. Access rated as lower priority in cognitive interviews in Bangladesh – MPNS7 likely to better capture experience. |
| 5 | Were you worried that your menstrual materials would allow blood to pass through to your outer garments? | High | X | X | Only item which captures the effectiveness of menstrual materials and lived experience of concern about leakage. Highly rated by girls in cognitive interviews. |
| 6 | Were you worried that your menstrual materials would move from place while you were wearing them? | Lower |  |  | Item is more applicable to those not using pads, partially captured by MPNS5, concerns about leakage. MPNS5 and 7 prioritised within this subscale. |
| 7 | Were you worried about how you would get more of your menstrual material if you ran out? | High | X |  | Preferred item to capture access to materials (between MPNS4 and 7). This captures both purchasing materials (for those purchasing their own) or asking their parents (for younger respondents who do not purchase/find their own materials). |
| 8 | Did you feel comfortable carrying spare menstrual materials with you outside your home? | Moderate |  |  | This item has a history of cross-loading as it bridges multiple locations (carrying from home to elsewhere). Better to remove for structural validity, however, participants often report high rates of *not* feeling comfortable transporting materials and this description is lost without this item. |
| 9 | Did you feel comfortable carrying menstrual materials to the place where you changed them? | Lower |  |  | Similar to MPNS8 this item often has issues with cross-loading or poorly loading on the school factor. The location for the item is not specified and may elicit different responses across the sample. |
| 10 | Did you feel comfortable storing your leftover or cleaned menstrual materials until your next period? | Low |  |  | Using this item in different languages we have encountered translation issues, and difficulties in clarity understanding this item. |
| 11 | Were you able to wash your hands when you wanted to? | Low |  |  | While this item asks about the last menstrual period, handwashing is not exclusive to menstruation and could be assessed as part of other measures. |
| 12 | Were you able to immediately dispose of your used menstrual materials? | Moderate |  |  | MPNS12 and 13 are often highly correlated (Khulna pilot polychoric correlation= .60). Immediate disposal may not always be the preferred mechanism; MPNS13 retained. |
| 13 | Were you able to dispose of your used materials in the way that you wanted to? | High | X | X | MPNS13 selected between the two disposal needs items. |
| 14 | Were you worried about where to dispose of your used menstrual materials? | Moderate |  |  | MPNS14 and 15 typically highly correlated (polychoric correlation=0.68 in Khulna pilot). MPNS13 captures use of preferred disposal locations, so privacy item was retained (MPNS15) |
| 15 | Were you concerned that others would see your used menstrual materials in the place you disposed of them? | High | X |  | Selected between MPNS14 and 15. Highest rated in sub-scale in cognitive interviews with girls. |
| 16 | Home - Were you able to change your menstrual materials when you wanted to? | High | X |  | Measures availability of changing locations at home. MPNS16 and 19 both capture this availability from different perspectives. MPNS16 retained in short form to balance positive and negative items and perceived to be more specific (ability to change) than MPNS 19 (worried would be unable to change). |
| 17 | Home - Were you satisfied with the place you used to change your menstrual materials? | High | X | X | Satisfaction can capture multiple dimensions of environment. Suggest item revision to incorporate example: *“For example, it was clean, safe, and had what you needed such as light or water”* |
| 18 | Home -Did you have a clean place to change your menstrual materials? | Lower |  |  | Captured by satisfaction. MPNS17 and 18 highly correlated in multiple samples. Cleanliness added as example to MPN17 with MPNS17 suggested for retention in short form. |
| 19 | Home - Were you worried that you would not be able to change your menstrual materials when you needed to? | Moderate |  |  | Retained item MPNS16 captures the availability/accessibility of spaces for changing materials, in favour of MPNS19. |
| 20 | Home - Were you worried that someone would see you while you were changing your menstrual materials? | High | X | X | Highly rated by girls in cognitive interviews. Privacy for changing in schools is included as a recommended indicator for national and global monitoring. |
| 21 | Home - Were you worried that someone would harm you while I you were changing your menstrual materials? | High | X |  | Applies to fewer participants, but incredibly important when it does apply. Safety in changing location at school included as recommended indicator for national and global monitoring. |
| 22 | Home - Were you worried that something else would harm you while you were changing your menstrual materials (e.g., animals, insects, unsafe structure) | Moderate |  |  | While safety of infrastructure and concerns about other harms are important, MPNS21 was prioritised. We considered combining MPNS21 and MPNS22 to capture safety more broadly, however we felt that retaining an understanding of concerns about safety from others was most important as reflective of concerns about gender-based violence. This granularity would be lost in a more general question. |
| 23 | School - Were you able to change your menstrual materials when you wanted to? | High | X | X | Highest rated in cognitive interviews with girls of ‘transport & school needs’ subscale. Captures accessibility and availability of changing spaces. |
| 24 | School - were you satisfied with the place you used to change your menstrual materials?<br>**Revised for testing to:** Were you satisfied with the places available at your school for changing your menstrual materials? | High | X |  | Matches with selected home-related item MPNS17. As for MPNS17, with added examples this can capture cleanliness as well as other required needs in the space. We have found that in multiple studies there are many participants who do not change their menstrual materials at school/work. This item was revised for the Khulna pilot and shortform to be relevant to those who do and don’t use facilities. Revised item: *“Were you satisfied with the places available at your school for changing your menstrual materials (for example it was clean, safe, and had what you needed such as light or water)?”* |
| 25 | School - Did you have a clean place to change your menstrual materials? | Moderate/<br>Essential |  |  | While this question is included as a recommended indicator for national and global monitoring, aligned with questions about the home environment MPNS24 capturing satisfaction (with expanded example text) is able to capture cleanliness along with broader needs. MPNS24 could be used to assess comparability with the national indicators and MPNS24 and 25 are highly correlated (polychoric correlation 0.80 in the Khulna pilot) |
| 26 | School - Were you worried that you would not be able to change your menstrual materials when you needed to? | Moderate |  |  | Both MPNS23 and 26 capture access to changing locations when needed/wanted. MPNS23 was selected, consistent with the item selected to capture experiences at home. Retaining balance between positively and negatively coded items. |
| 27 | School - Were you worried that someone would see you while you were changing your menstrual materials? | Essential | X | X | Recommended indicator for national and global monitoring, high priority for girls who do change menstrual materials at school. |
| 28 | School - Were you worried that someone would harm you while you were changing your menstrual materials? | Essential | X |  | While less often endorsed, highlights important safety concerns and perceptions of gender based violence when reported. |
|  | **Reuse items** |  |  |  |  |
| 29 | Did you have enough water to soak or wash your menstrual materials? | High | X | X | For shortform, proposed to combine MPNS29 and MPNS32 since the use of soap requires water. Revised item for shortform *“Did you have enough water and soap to wash your menstrual materials?”* |
| 30 | Did you have access to a basin or bucket to soak or wash your menstrual materials whenever you needed it? | Low |  |  | Having enough water and soap accessible often requires having a washing vessel. MPNS29/32 were prioritised over this item. |
| 31 | Were you able to wash your menstrual materials whenever you wanted to? | High | X |  | Rated as a very important item for reusing materials in cognitive interviews that included girls with experience using reusable materials. Captures the accessibility/availability of washing and potential restriction on time. |
| 32 | Did you have enough soap to wash your menstrual materials? | High |  |  | MPNS29 and MPNS32 combined into a single item for the short form. |
| 33 | Were you able to dry your materials when you wanted to? | Moderate |  |  | This is the only item that captures satisfaction with the experience of drying, rather than worries/concerns. The availability of drying is often impacted by weather and privacy concerns, which are captured by MPNS35 and MPNS36. |
| 34 | Were you worried that someone would see you while you were washing your menstrual materials? | Moderate |  |  | While highly relevant, MPNS31 highlights the availability of washing, which is likely to be impacted by privacy. Reuse items are typically highly correlated across samples. MPNS35 and 36 were preferred. |
| 35 | Were you worried that your menstrual materials would not be dry when you needed them? | High | X |  | This item is impacted by both drying practices (e.g., drying under other fabric to hide menstrual materials) but also seasonal weather (e.g., wet seasons). Impacts material availability and hygiene if wet materials are reused. |
| 36 | Were you worried that others would see your menstrual materials while they were drying? | High | X | X | Highly rated by girls in cognitive interviews with experience of reusing materials. Impacts drying hygiene and experience. |

### Short form dimensionality

Dimensionality of the short form was assessed in the Khulna Pilot, Bangladesh. The original four-factor structure, and two reuse factors, remained an acceptable fit. In testing in the Khulna pilot sample, EFA was undertaken and further indicated that the original four-factor structure offered the best sub-scale solution. Single and two-factor structures did not offer better fit for the data and demonstrated poor factor loading for school-related items and material reliability concerns.

Structural validity was then replicated in the Khulna Cohort Baseline, Bangladesh and Soroti, Uganda populations. CFA findings for all three datasets are presented in Table 3. Despite the small sample size among the Bangladesh samples including girls who changed their menstrual materials at school, the four-factor structure was an acceptable fit, achieving a good fit in the original Soroti, Uganda data with a more substantial sample.

**Table 3.**
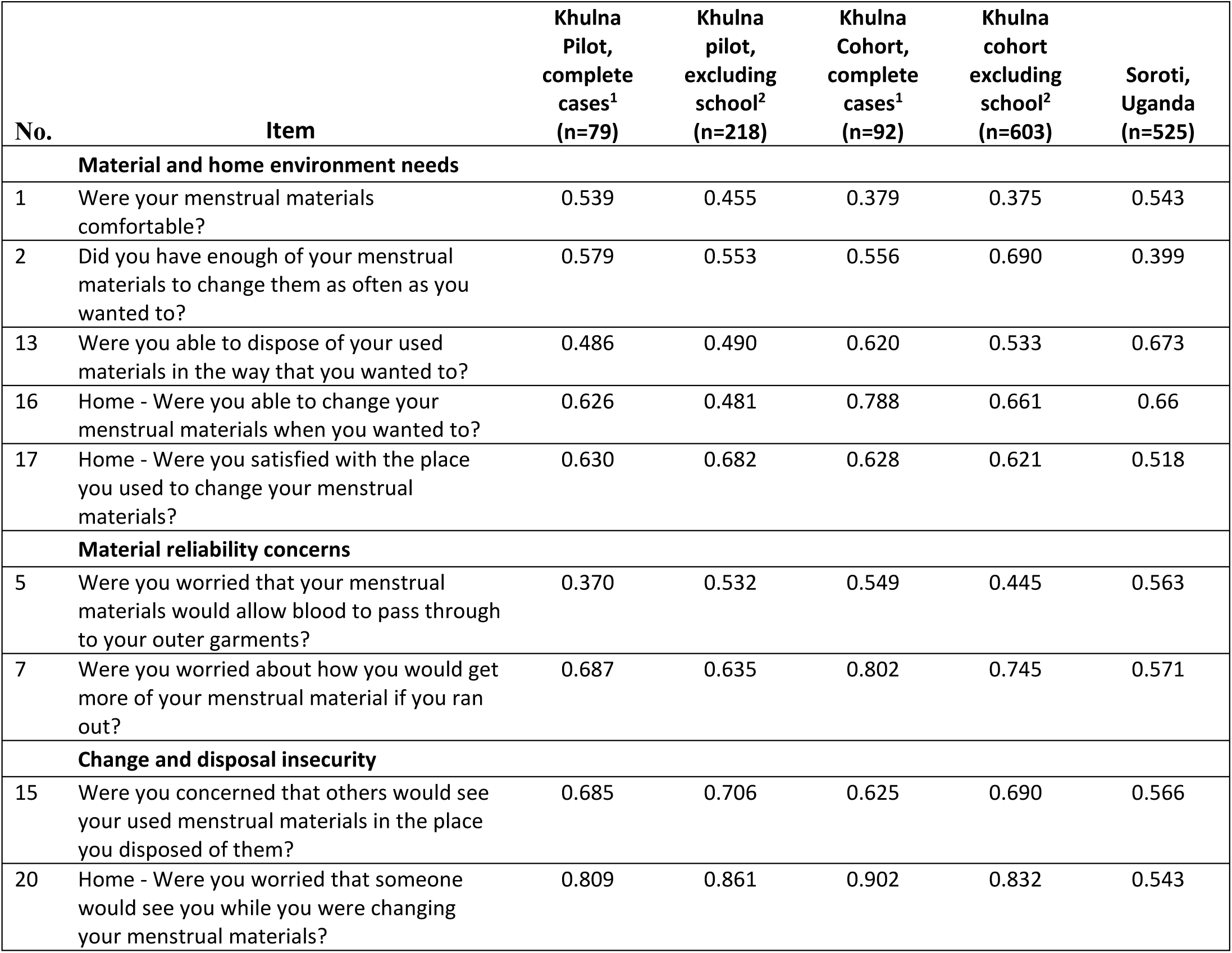
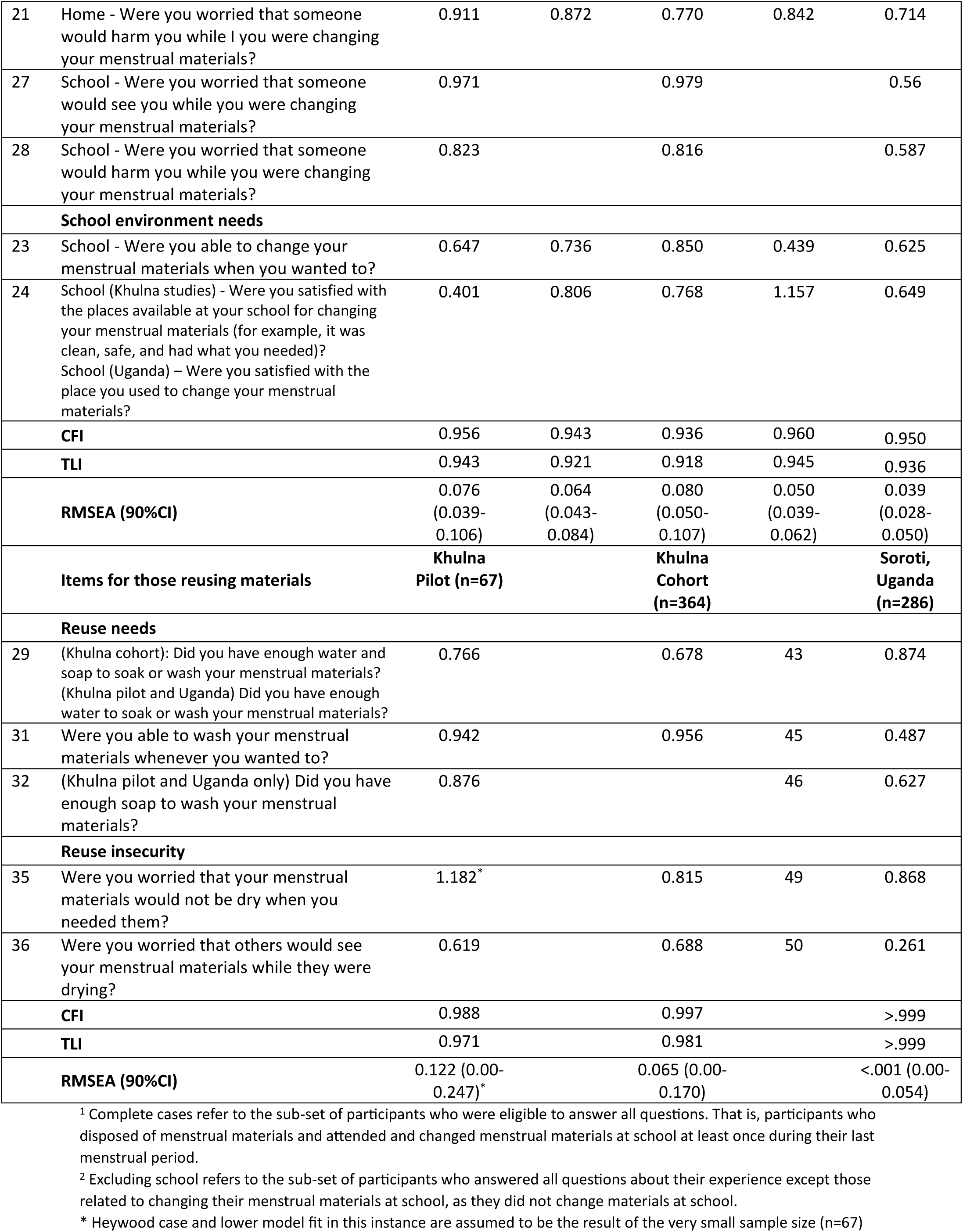
Dimensionality of the MPNS-SF in all three datasets, with factor loadings presented by sub-scale.

For girls who did not change their menstrual materials at school, and thus did not answer MPN27 and MPN28, the four factors were an acceptable fit for the data. However, we note that a Heywood case was observed in the cohort baseline data which excluded these items (see Table 3). In exploring this case, we found that both MPN23 and MPN24 have low bivariate correlations with other non-school related items in the measure (all polychoric correlations <.15 in both the pilot and cohort samples) including some 0 bivariate relationships. They have correlations with MPN27 and MPN28 (polychoric correlations .22-.32 across both samples) which in turn have meaningful correlations with other items across the scale. When removing MPN27 and MPN28 to test the factor structure in girls not changing menstrual materials at school, it is likely these zero-correlations (and the low correlation between the remaining school subscale and other factors (correlations=0.07-0.15) results in a Heywood case. It is consistent with our expectations of the measure that girls’ satisfaction with the available school facilities will not have a strong relationship to their satisfaction with their materials or home environment, while insecurities in the home and school environment have a closer relationship demonstrated by loading on the same factor. The sample excluding girls who change at school in this analysis also means that girls’ reports about their satisfaction with their disposal experience captures only the home environment, further minimising relationships between MPN23/24 and other scale items. As such we did not interpret the Heywood case as indicating model misspecification.[29] However, this finding suggests that the sub-scale structure is less stable when the full set of items is not included due to eligibility constraints.

### Rapid form dimensionality

Items in the rapid form prioritised breadth. As a result, most factors were only represented by one or two items; with single item factors unable to be included in CFA for model fit. As we hypothesised based on findings from the full and short form MPNS, a single-factor solution was not an acceptable fit for the data (excluding reuse items) in the Khulna pilot dataset (CFI = 0.897, TLI = 0.828, RMSEA 0.131, 90%CI 0.058-0.205). A two-factor solution offered the best fit in the Khulna Pilot but did not reach thresholds of acceptable fit for all metrics, and this was replicated in the other two datasets (see Table 4). School-related items loaded poorly on the two factors. However, we note the small sample size to test the full set of rapid-form items in the Khulna pilot and cohort datasets given the limited number of girls who changed their menstrual materials at school. In the datasets including the larger number of participants who did not change their menstrual materials at school, remaining school-related items fit poorly with the factor structure and compromised stability of the model.

**Table 4.**
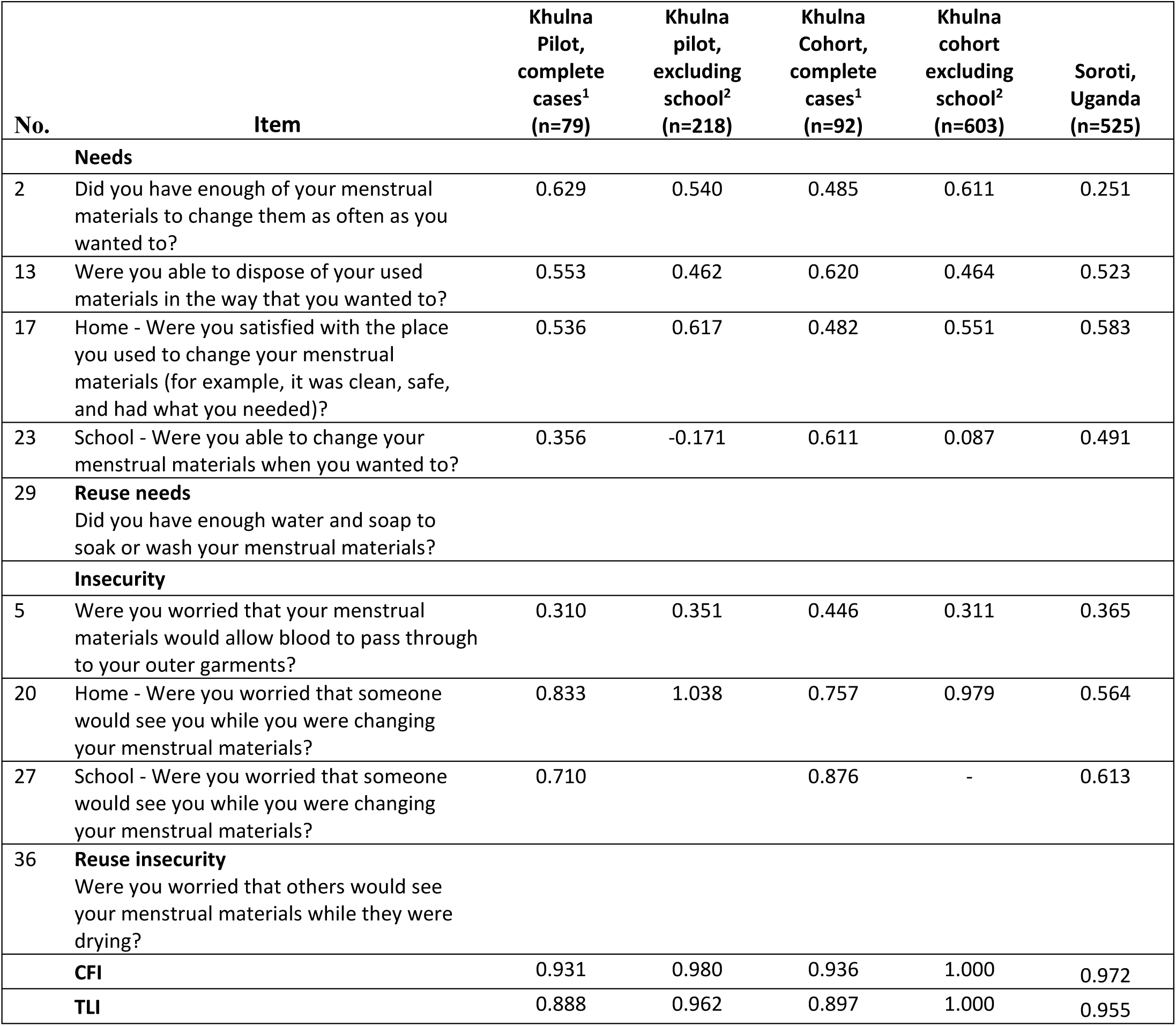
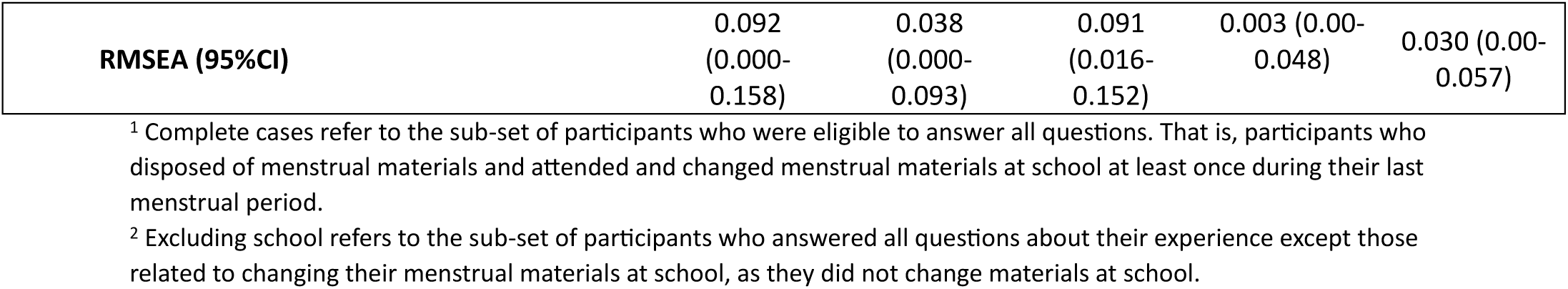
Dimensionality of the MPNS-R across datasets, with factor loadings presented by sub-scale.

### Validity and internal consistency

Mean, standard deviation, internal consistency and validity tests are presented in Table 5 for the respective MPNS-36, MPNS-SF and MPNS-R total scales. Findings for MPNS-SF sub-scales are presented in Supplementary Materials 3. The MPNS-SF total scales displayed adequate internal consistency across datasets, with sub-scales largely displaying strong reliability particularly when tested using α for ordinal items. Two-item sub-scales such as ‘material reliability concerns’ exhibited poorer reliability as expected for the small number of items. The MPNS-R had acceptable ordinal internal consistency in Bangladesh, but poorer reliability in the Soroti, Uganda sample.

**Table 5.** MPNS-36, MPNS-SF and MPNS-R total scale scores, internal consistency, and associations with hypothesised correlates.

| | Mean | SD | $\alpha$ | $\alpha_{\text{ordinal}}$ | DASS<br>r (p-value)<sup>1</sup> | Did not miss<br>school due to last<br>period<br>OR (95%CI) | Did not<br>experience<br>difficulty<br>participating in<br>class due to last<br>period<br>OR (95%CI) | Very confident<br>managing<br>menstruation at<br>home<br>OR (95%CI) | Confident<br>managing<br>menstruation at<br>school<br>OR (95%CI) |
| --- | --- | --- | --- | --- | --- | --- | --- | --- | --- |
| **KHULNA PILOT** |  |  |  |  |  |  |  |  |  |
| MPNS-36 total | 2.16 | 0.37 | 0.86 | 0.92 | -0.34 (p<.001) | 1.22 (0.66-2.26) | 2.72 (1.37-2.39) | 1.13 (0.58-2.16) | 3.94 (2.01-7.73) |
| MPNS-SF total | 2.21 | 0.39 | 0.78 | 0.88 | -0.28 (p<.001) | 1.07 (0.60-1.90) | 2.56 (1.35-4.86) | 1.40 (0.75-2.60) | 2.91 (1.57-5.39) |
| MPNS-R total | 2.14 | 0.45 | 0.59 | 0.74 | -0.32 (p<.001) | 1.09 (0.66-1.83) | 2.36 (1.33-4.19) | 1.26 (0.73-2.16) | 2.32 (1.35-3.98) |
| **KHULNA COHORT** |  |  |  |  |  |  |  |  |  |
| MPNS-SF total | 2.08 | 0.44 | 0.78 | 0.87 | -0.36 (p<.001) | 1.88 (1.37-2.57) | 4.80 (3.36-6.86) | 2.40 (1.66-3.47) | 1.58 (1.17-2.14) |
| MPNS-R total | 1.99 | 0.49 | 0.60 | 0.72 | -0.32 (p<.001) | 1.77 (1.33-2.36) | 3.65 (2.65-5.01) | 2.36 (1.68-3.30) | 1.54 (1.17-2.03) |
| **SOROTI** |  |  |  |  |  |  |  | **Confident**<br>**managing**<br>**menstruation at**<br>**home**<br>**OR (95%CI)** |  |
| MPNS-36 total <sup>2</sup> | 1.82 | 0.37 | 0.77 | 0.82 | -0.11 (p=.013) | 2.62 (1.52-4.50) | - | 4.09 (2.52-7.06) | 4.22 (2.52-7.06) |
| MPNS-SF total | 1.79 | 0.40 | 0.64 | 0.72 | -0.12 (p=.005) | 2.42 (1.48-3.97) | - | 3.09 (1.75-5.45) | 4.02 (2.50-6.46) |
| MPNS-R total | 1.79 | 0.47 | 0.43 | 0.48 | -0.04 (p=.310) | 1.48 (0.98-2.22) | - | 2.27 (1.41-3.65) | 3.46 (2.31-5.19) |

<sup>1</sup> Khulna Pilot: Only half of the sample was administered the DASS, associations presented here are for this half of the sample (n=164 of 313 menstruating girls) | Khulna Cohort and Soroti data. <sup>2</sup> Findings from the original study report[3], provided here for ease of comparison

Short and rapid forms of the measure were found to demonstrate hypothesised relationships with school participation, and confidence managing menstruation across datasets. For each reduction of the scale, the strength of these associations was attenuated.

## Discussion

Measuring adolescent girls’ experiences of managing their menstrual bleeding is essential to capture whether their menstrual health needs are being met, test associations between this experience and broader health and wellbeing outcomes in research, and to understand the impacts of menstrual health interventions. This study developed short and rapid forms of the MPNS-36, as requested by stakeholders. The MPNS-SF offers an 18-item measure, including 14 items if respondents do not reuse any menstrual materials, with 4 items specific to laundering. Two items specific to disposal are relevant to those who disposed of single-use or reusable materials during their last period. The MPNS-R includes 9 items, 7 for respondents who did not reuse menstrual materials. The MPNS-SF offers consistent sub-scales to the original measure, with acceptable dimensionality, internal consistency, and strong validity. The MPNS-R reduces participant burden further whilst sacrificing structural validity and attenuating the relationship between the measure and hypothesised correlates. All three forms offer strong face validity, have been well understood by adolescent populations in cognitive interviews, and a high level of interpretability such that both individual items and scale scores offer easily understandable insights into girls’ menstrual experiences, needs and relationships with other outcomes.

Our findings highlight that girls’ experiences of managing menstrual bleeding are multi-dimensional, driven by the diverse practices required and environments in which menstruation is managed. Despite halving the number of items for the short form, the four and two factor structure remained the best fit for the data across all three available datasets. Consistent with past studies in Uganda,[3, 6] experiences at home and at school differed substantially in the samples in Bangladesh. However also consistent with past application of the measure among adolescents, girls’ insecurities, that is worries about availability, privacy, and safety, regarding spaces for menstrual management loaded on a single factor across locations. These concerns may have more unifying drivers across locations such as girls’ internalised stigma regarding menstruation or trait anxiety.

In selecting short-form items, we prioritised maintaining content validity and the breadth of practices captured. However, we note that items capturing experiences of transporting and storing menstrual materials were lost in moving from the original 36-item to short and rapid measures. Experiences of transporting items have consistently presented issues in cross-loading given they bridge multiple environments (e.g., transporting materials from home to school)[3, 6]. In data collected in the Khulna pilot survey, we note that almost half of girls reported they never felt comfortable carrying spare menstrual materials outside their home. Future research must remain sensitive to this important, but often neglected, challenge.

We found that capturing experiences across the breadth of menstrual practice offers the strongest correlation with hypothesised related constructs and impacts of menstrual experience. This was observed across the original, short, and rapid forms of the measure wherein total scores exhibited stronger relationships with hypothesised correlates than sub-scales capturing only one dimension of blood management experience. The finding supports our hypothesis that capturing menstrual experience requires the use of multiple-item measures, and aligns with qualitative research which has consistently highlighted the diversity of menstrual management challenges that impact on women’s and girls’ lives.[16, 30, 31]

### Strengths and limitations

We triangulated insights from implementation of the MPNS, the perspectives of adolescent girls in Bangladesh, and advances in menstrual health research guidance on monitoring [17] to develop the MPNS short and rapid forms. A data-driven approach to shortening the scale would likely have yielded greater model fit of sub-scales but would reduce the breadth of practices and thus validity of the measure. A strength of the MPNS remains its development drawing on synthesis of qualitative research of women’s and girls’ experiences of menstruation across low-and-middle income country settings.[3, 16] The variety in menstrual practices that adolescent girls employ means not all MPNS items are relevant to all respondents. Non-applicable items reduce the sample sizes available for undertaking tests of dimensionality with complete data. In our Bangladesh samples where many girls do not change their materials at school meant restricted samples were available to test dimensionality for all items. While measures are emerging for capturing menstrual experiences in different populations,[32, 33] we did not have other measures of adolescent menstrual experiences against which to test convergent or divergent validity.

### Implications for research and practice

Caring for the body during menstruation in a way that meets individual needs for hygiene, comfort, privacy, and safety is a requirement for menstrual health, and objective of many menstrual health policies and programs.[13, 34] The MPNS-SF and MPNS-R offer concise measures to capture this construct. Based on the findings of measure development, the MPNS-36 still offers the greatest nuance in describing population needs and explanatory power in investigating hypothesised correlates. The MPNS-36 may be best suited for needs assessment in a new population. The MPNS-SF offers a reliable and valid measure, with sub-scales matching the original, associations with hypothesised correlates, and good internal consistency. The measure maintains a balance in practices and captures both having needs satisfactorily met and insecurities related to menstrual management. It also offers minor modifications to improve the applicability of some items in the school environment. We recommend that the MPNS-SF be used when a shorter measure is needed such as in research and intervention evaluation where survey length is restricted, or multiple constructs must be accommodated. Finally, the MPNS-R provides a restricted set of questions with brief survey duration. In shortening the measure further, the MPNS-R no longer offers reliable sub-scales, with multiple original sub-scales represented by a single item. However, the tool retains breadth in capturing experiences of menstrual practices and environments and is preferable to selecting sub-scales of the original measure or ad-hoc item selection which results in incomparable data across studies. We recommend the rapid form is used where menstrual health is not the primary area of focus but included as part of studies of broader water, sanitation, and hygiene (WASH) or sexual and reproductive health (SRH).

Future longitudinal research using the MPNS measures will test their predictive validity and the impact of menstrual experiences on individuals’ lives. Subsequent waves of the AMEHC Bangladesh study will offer these opportunities,[20] and more cohort studies will be needed across contexts. Trials using the MPNS measures as primary or secondary outcomes will provide insights into the sensitivity of the measure to change and use for evaluation.[8, 12] Finally, future research should investigate the validity of the shorter MPNS forms in new cultural contexts, languages and age groups. As found previously,[6] the dimensionality of the MPNS-36 differs for adult women, and we would hypothesise similar differences for the short form. Improved availability of high-quality measures for menstrual health research and practice will strengthen the evidence base and aid comparability across studies.[1, 35, 36]

### Ethical approvals

#### Khulna Pilot and Cohort, Bangladesh

The study received ethical approval from the Alfred Hospital Ethics Committee, Melbourne, Australia (369/22) and the Institutional Review Board of BRAC James P Grant School of Public Health, BRAC University (IRB-06 July 22-024), Bangladesh. National and district-level education offices provided endorsement for the study.

#### Soroti, Uganda

Publicly available de-identified data is available at https://osf.io/qshkc/. The original study was approved by Johns Hopkins School of Public Health Institutional Review Board (IRB approval no: 00009073) and the Mildmay Uganda Research Ethics Committee (MUREC) (approval ref: 0212–2018). The Uganda National Council for Science and Technology (UNCST) approved the study (ref: SS279ES).

## Author contributions (CRediT statement)

Conceptualization: JH

Methodology: JH

Investigation: JH, PA, EH, AH, AJ, MTH, TJ, AM, NSZ, MKA, AK, LD, AZ, AK, CS, LB

Data curation: JH, AH, MKA, AM, SA

Formal Analysis: JH

Writing – Original draft: JH

Writing – review & editing: All authors

Supervision: JH, MTH, PSA, CS

Project Administration: JH, MTH, CS

Funding acquisition: JH, PSA

All authors have approved the final manuscript.

## Funding

Development of the MPNS short form was funded by The Case for Her, the Reckitt Global Hygiene Institute (RGHI), and supported by funding from the National Health and Medical Research Council of Australia (NHMRC) (GNT2008600). The AMEHC study is funded by the National Health and Medical Research Council of Australia (NHMRC) (GNT2004222 and GNT2008600), and the Reckitt Global Hygiene Institute (RGHI). The views expressed are those of the authors and not necessarily those of RGHI. J Hennegan is supported by an RGHI Fellowship and NHMRC Investigator Grant (GNT2008600). P Azzopardi is supported by a NHMRC Investigator Grant GNT2008574.

The authors gratefully acknowledge the contribution to this work of the Victorian Operational Infrastructure Support Program received by the Burnet Institute.

## Data Availability

Khulna Pilot and Baseline. De-identified data relevant to this manuscript are available in an open access repository at https://osf.io/uh9z8/.
Soroti, Uganda: Data are available in a public, open access repository. Deidentified data are available at https://osf.io/qshkc/.

## Acknowledgements

We thank the many field research assistants who administered the surveys, without whom the study would not have been possible. Our deepest thanks to the participating schools, families, and girls.

We are grateful for input from the AMEHC study National Advisory Committee, and the Directorate of Secondary and Higher Education in Bangladesh.

## Supplementary Materials 1

Cognitive Interviews. Selection of participants’ explanations of their responses.

|  | MPNS Item | Participant response |
| --- | --- | --- |
| 1 | Were your menstrual materials comfortable? | *Sometimes* - the pads are uncomfortable and sometimes itchy. (CI5)<br>*Sometime*-because she uses pads and cloths, but when she uses cloth it's uncomfortable. (CI3)<br>*Always*. She is comfortable with pads and cloth. (CI3) |
| 2 | Did you have enough of your menstrual materials to change them as often as you wanted to? | *Sometimes*. Pad is not available always. (CI2)<br>*Always*. I usually change cloth after 30 minutes (CI3)<br>*Always*. As they always have enough pads stored in their house. (CI5) |
| 3 | Were you satisfied with the cleanliness of your menstrual materials? | *Sometimes*-I can't always clean my cloths when I'm at school. (CI1)<br>*Always*-I can change my pads after every six hours. (CI3)<br>*Always*-I clean my used cloths with Savlon and powder soap. (CI3) |
| 4 | Could you get more of your menstrual materials when you needed to? | *Always*. She says that when she runs out of pads, she tells her mother. Then her mother would have brought pad for her. (CI7)<br>*Most of the time*. My mother always buy my pads but when it finished I used cloths. (CI3) |
| 5 | Were you worried that your menstrual materials would allow blood to pass through to your outer garments? | *Never*. She says her bleeding was very light during her last period. Moreover, she was wearing pad properly. (CI7)<br>*Sometimes* - mentioned getting worried on heavy days but not always. (CI6)<br>*Most of the time* –I am always tensed about it. (CI1)<br>*Always*, as they are constantly worried about leakage due to heavy bleeding. (CI5) |
| 6 | Were you worried that your menstrual materials would move from place while you were wearing them? | *Never*. She is very confident that her menstrual pad will not dislodge. It never happened. (CI4)<br>*Sometimes*. She says that since she used the pad for the first time, she sometimes felt like the pad was slipping out of place. (CI7)<br>*Most of the time*-cause most of the time I use cloths. (CI1) |
| 7 | Were you worried about how you would get more of your menstrual material if you ran out? | *Never*. Says that her mom always brings her menstrual material. Her mother even brought the menstrual material before it was over. (CI4)<br>*Never*. Because they always have pads in their house. (CI6) |
| 8 | Did you feel comfortable carrying spare menstrual materials with you outside your home? | *Most of the time* - I keep them in a separate pocket which no one opens (CI8)<br>*Sometimes*. When we go relatives house we can't carry, if brother or father see us to carry pads, we feel awkward. (CI9) |
| 9 | Did you feel comfortable carrying menstrual materials to the place where you changed them? | *Sometimes*-we carry it with us to bathroom hiding under the scarf. (CI2)<br>*Sometimes* because she also carries the pads in her sleeves or covers them with a scarf (CI5) |
| 10 | Did you feel comfortable storing [keeping] your leftover or cleaned menstrual materials until your next period? | *Always*. She says she keeps the unused pads for next time. But she keeps it hidden. (CI7)<br>*Always*, as they use pads and they store their leftover pads in the drawer for their next cycle. (CI5) |
| 11 | Were you able to wash your hands when you wanted to? | *Always*-Both girl clean their hands when they touch menstrual materials. (CI2)<br>*Always*. because they have enough supply of water and facilities. (CI5) |
| 12 | Were you able to immediately dispose of your used menstrual materials? | *Most of the time.* She says she throws her used menstrual pads in the dustbin. But it is not always possible to throw it in the dustbin immediately. Because the dustbin was the outside of her house. (CI7)<br>*Most of the time-* I can't when guests are there. (CI3) |
| 13 | Were you able to dispose of your used materials in the way that you wanted to? | *Sometimes-*we store our used pads altogether and throw away when menstruation ends. (CI1)<br>*Always.* Yes, I have always thrown it the way I wanted. We have a dustbin in our house. Always left there. (CI7) |
| 14 | Were you worried about where to dispose of your used menstrual materials? | *Never.* She said that she throws her menstrual materials in the dustbin at home. During throw the pad she wraps it well in paper. Then the garbage men come and take away the garbage. So, she didn't have to worry. (CI4)<br>*Sometimes -*because, when they are outside of the house they get worried about where to dispose of the used pads. (CI5) |
| 15 | Were you concerned that others would see your used menstrual materials in the place you disposed of them? | *Never.* She says that used menstrual pads are wrapped in paper or plastic... so no one can see (CI4)<br>*Some of the time.* Because she disposes of it in a dumpster near her house, where neighbours can see the materials. (CI6)<br>*Always-* and said that they are worried about disposing of the materials. Because their neighbours or others might see them while disposing of them and ask questions. (CI5) |
| 16 | Home - Were you able to change your menstrual materials when you wanted to? | *Sometimes* because they are worried that their family members might bother them by asking questions about pads if they (male members) see them. So, when the male members are near the bathroom they don't change. (CI5)<br>*Always.* Because they have a separate bathroom in their house. (CI3) |
| 17 | Home - Were you satisfied with the place you used to change your menstrual materials? | *Always-*yes, we can change our pads in our room and also in bathroom (CI1) |
| 18 | Home -Did you have a clean place to change your menstrual materials? | *Always.* A clean place where no dirt is present (CI8) |
| 19 | Home - Were you worried that you would not be able to change your menstrual materials when you needed to? | *Never.* She also feels that she never has to worry about changing pads while at home. She had enough opportunity to change pads at home. (CI7)<br>*Never,* we have two separate bathrooms at our home. My parents use one and we use another one. So I can change whenever I want... But girls who live in the slum area are in risk, anyone can see them (as they use open toilet usually) (CI7) |
| 20 | Home - Were you worried that someone would see you while you were changing your menstrual materials? | *Sometimes.* She says that sometimes her mother sees her while changing pads. But she is not worried about that. But she is also careful when changing pads so that no one sees her. (CI4)<br>*Most of the time and always.* They said that their family members might see them while changing pads/ taking the pads with them to the bathroom/disposing of them. (CI5) |
| 21 | Home - Were you worried that someone would harm you while I you were changing your menstrual materials? | *Never.* They asked, who is gonna hurt them? And how? (CI6) |
| 22 | Home - Were you worried that something else would harm you while you were changing your menstrual materials (e.g., animals, insects, unsafe structure) | *Never.* She was very surprised to read the question. Said she was never worried about this. (CI4)<br>*Never.* They change their pads in the bathroom which is clean and safe. (CI6) |
| 23 | School - Were you able to change your menstrual materials when you wanted to? | *Sometimes*- because they felt shy to ask permission from their male teachers to go to the bathroom with a friend to change their pads. (CI5)<br>*Most of the time*-can't go to the bathroom unless the teacher gives permission. (CI3) |
| 24 | School – Were you satisfied with the place you used to change your menstrual materials? | *Never*-Sometimes we can't find dustbin in bathroom (CI1)<br>*Sometimes*- Our school's bathroom is clean. (CI2)<br>*Most of the time*. We have a bin (balti) which get filled sometimes, then we cannot dump pads (CI8) |
| 25 | School - Did you have a clean place to change your menstrual materials? | *Sometimes* because usually, their bathroom is clean but it gets muddy as many students use it. (CI5)<br>*Most of the time*-our school's bathroom is neat and clean. (CI1)<br>*Most of the time*-Our school's bathroom is clean and it has dustbin box. (CI3) |
| 26 | School - Were you worried that you would not be able to change your menstrual materials when you needed to? | *Sometimes*. We can go to the toilet. But they can see while carrying the pads to the toilet underarm. (CI8)<br>*Never*. She thinks that anyone can change their menstrual materials as needed at school. She also said that their school bathroom is very good and clean. (CI4) |
| 27 | School - Were you worried that someone would see you while you were changing your menstrual materials? | *Never*. because they lock the door from the inside of the bathroom, so no one can see them. (CI6)<br>*Never*- Our bathroom is separated from boys bathroom. (CI1)<br>*Most of the time* because they are worried about getting bullied by the students both female and male, if other students see pads with them. (CI5) |
| 28 | School - Were you worried that someone would harm you while you were changing your menstrual materials? | *Never*. She said that their school is a girl's school. No boys are allowed here. Therefore, she did not think that someone could be harmed during the change of menstrual material. (CI4) |
| 29 | Did you have enough water to soak or wash your menstrual materials? | *Always*. We have tubewell so there is no problem with water (CI2) |
| 30 | Did you have access to a basin or bucket to soak or wash your menstrual materials whenever you needed it? | *Always*-Both girls have separate pot and mug for menstrual cloths cleaning. (CI3) |
| 31 | Were you able to wash your menstrual materials whenever you wanted to? | *Most of the time*-when we are in Madrasa or outside of home then we can't do this. (CI2)<br>*Most of the time*. I have menstrual cramps, so I could not do it always if I want to do so. I cannot stand when I have cramps (CI8) |
| 32 | Did you have enough soap to wash your menstrual materials? | *Most of the time*. We do not get soap to clean menstrual pads at madrasa always. (CI8) |
| 33 | Were you able to dry your materials when you wanted to? | *Most of the time*-I can't dry clothes in rainy season, it's takes time. (CI2)<br>*Most of the time*. If I use cloths I hide them with other cloths to dry. Then we cannot dry cloths properly always. (CI8)<br>*Always*-I can dry clothes always even by fan. (CI2) |
| 34 | Were you worried that someone would see you while I you were washing your menstrual materials? | *Never*-No one ever sees and we also wash our cloth in bathroom. (CI2)<br>*Most of the time*. Males can see us at home, as we live jointly. (CI9) |
| 35 | Were you worried that your menstrual materials would not be dry when you needed them? | *Sometimes*-When there is no pad, then I got worried. (CI3) |
| 36 | Were you worried that others would see your menstrual materials while they were drying? | *Never*-we dry our clothes with fan air or in backside of stand or wardrobe. (CI2)<br>*Sometimes*. My brother can see it at home, as we dry it behind a curtain, curtain can move anytime (CI9) |

## Supplementary Materials 2: Revalidation of the original MPNS-36 in Bangladesh (Khulna)

### Dimensionality

Table 1 presents item responses, and factor loadings for confirmatory factor analysis (CFA) undertaken to test the performance of the original MPNS-36 factor structure in our *Bangla* version of the measure in Khulna, Bangladesh. The original MPNS-36 factor structure showed strong performance. Despite only 79 cases will full data for items related to changing menstrual materials at school, the original 4-factor structure for participants disposing of materials performed well (CFI=0.924, TLI=0.927, RMSEA=0.075, 90%CI=0.060-0.090). Nearly all factor loadings exceeded 0.5, with poorer fitting items including items related to storing menstrual materials (MPN10), concerns materials would move from place (MPN6) and MPN8 and MPN9 capturing concerns about transporting materials; these items have historically exhibited challenges as they are relevant across locations (not only at home or at school). When removing the items related to changing menstrual materials at schools, relative fit statistics indicated a poorer relative fit for the 4-factor model (CFI=0.886, TLI=0.872), but the model still achieved acceptable absolute fit (RMSEA=0.067, 90%CI 0.058-0.067). Consistent with past studies, the items capturing experiences of washing and drying absorbents showed strong fit with a two-factor model, and performed well when assessed alongside the 4-factors for those changing in school (see Table 1 below).

**Table 1.**
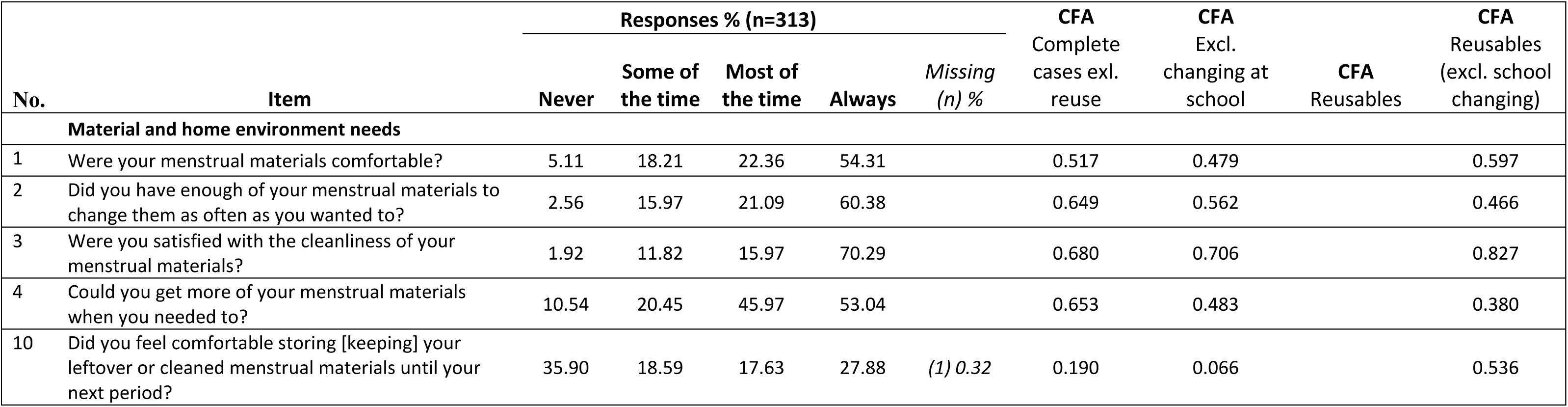
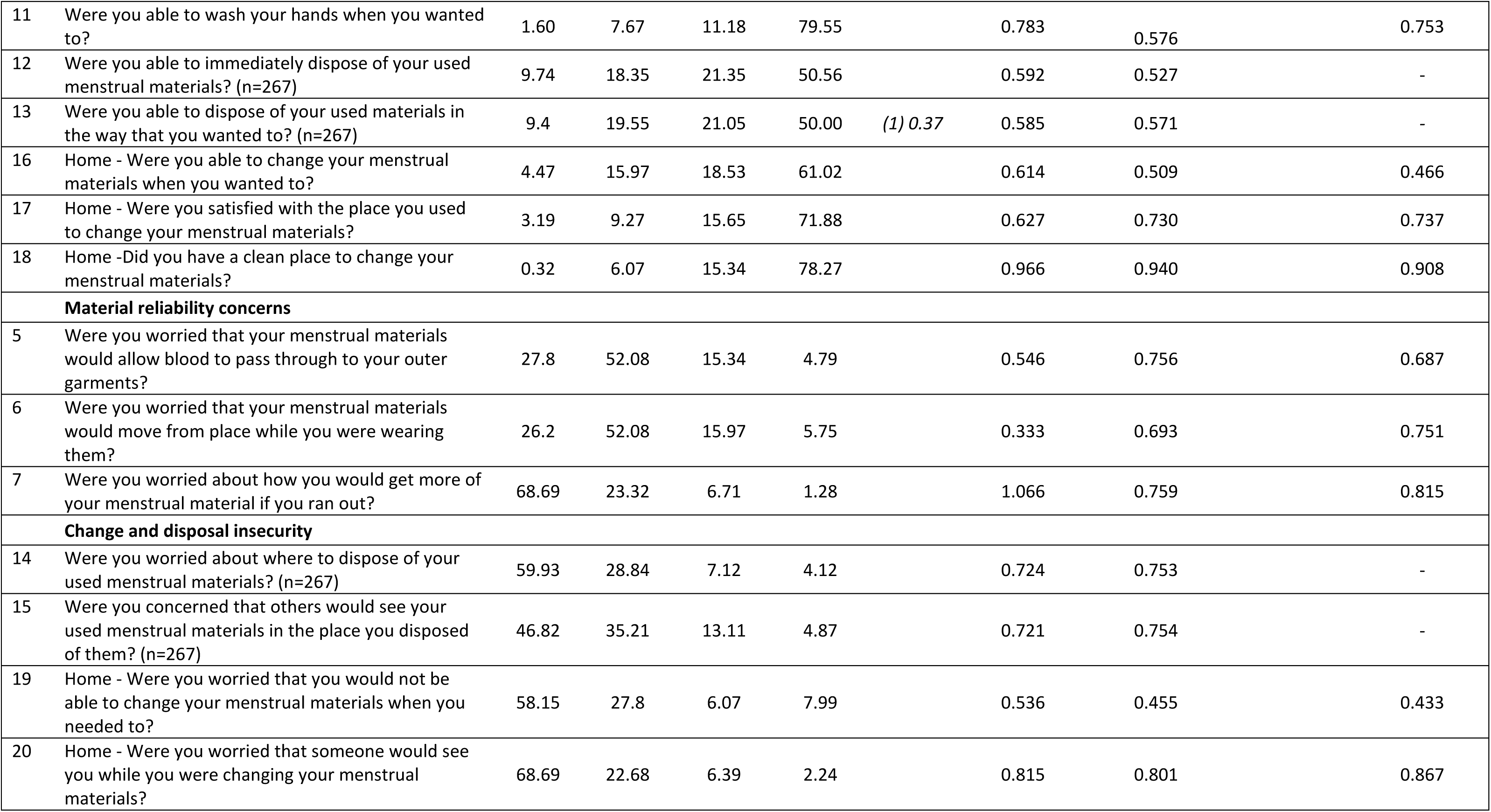
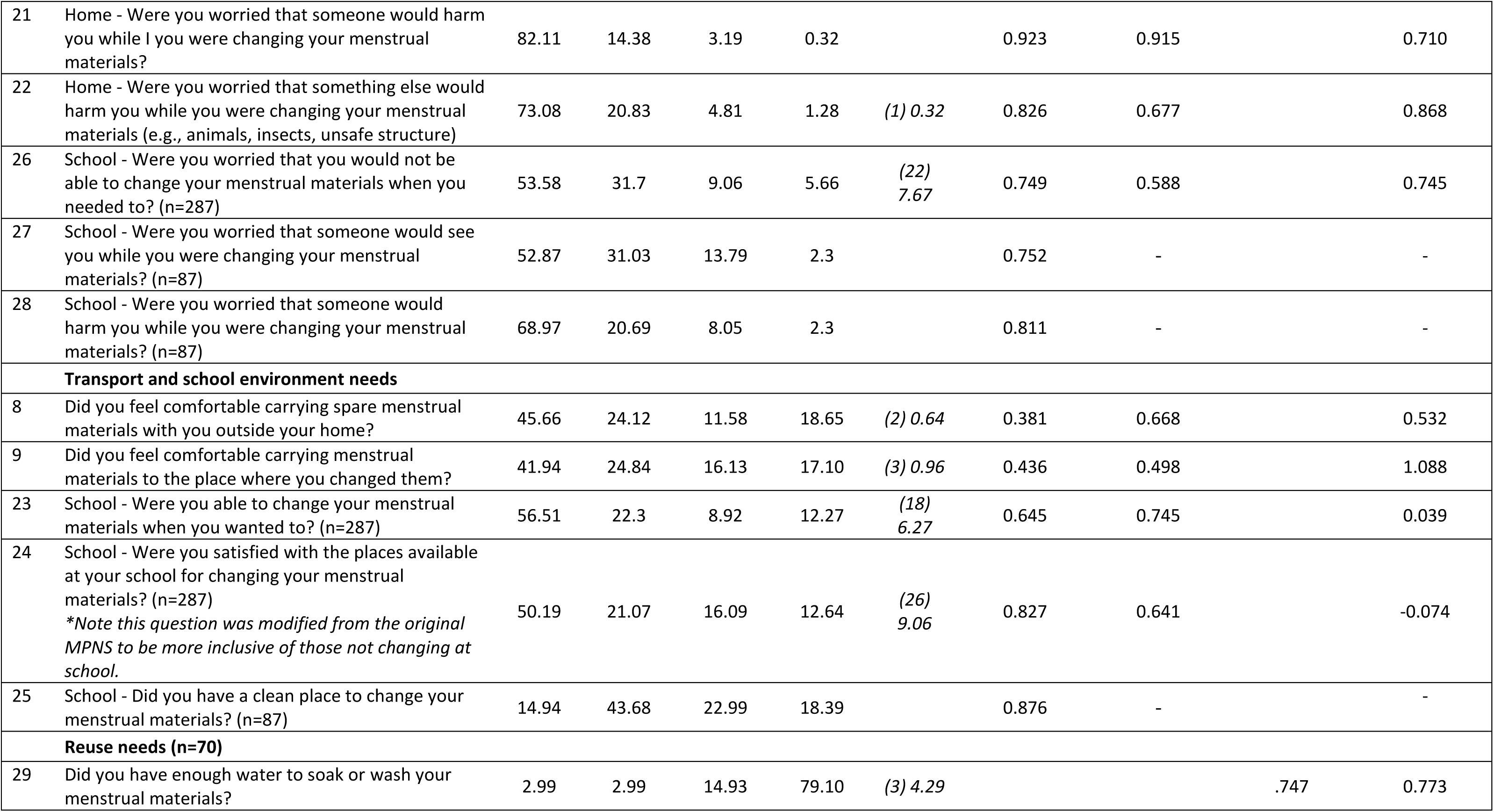
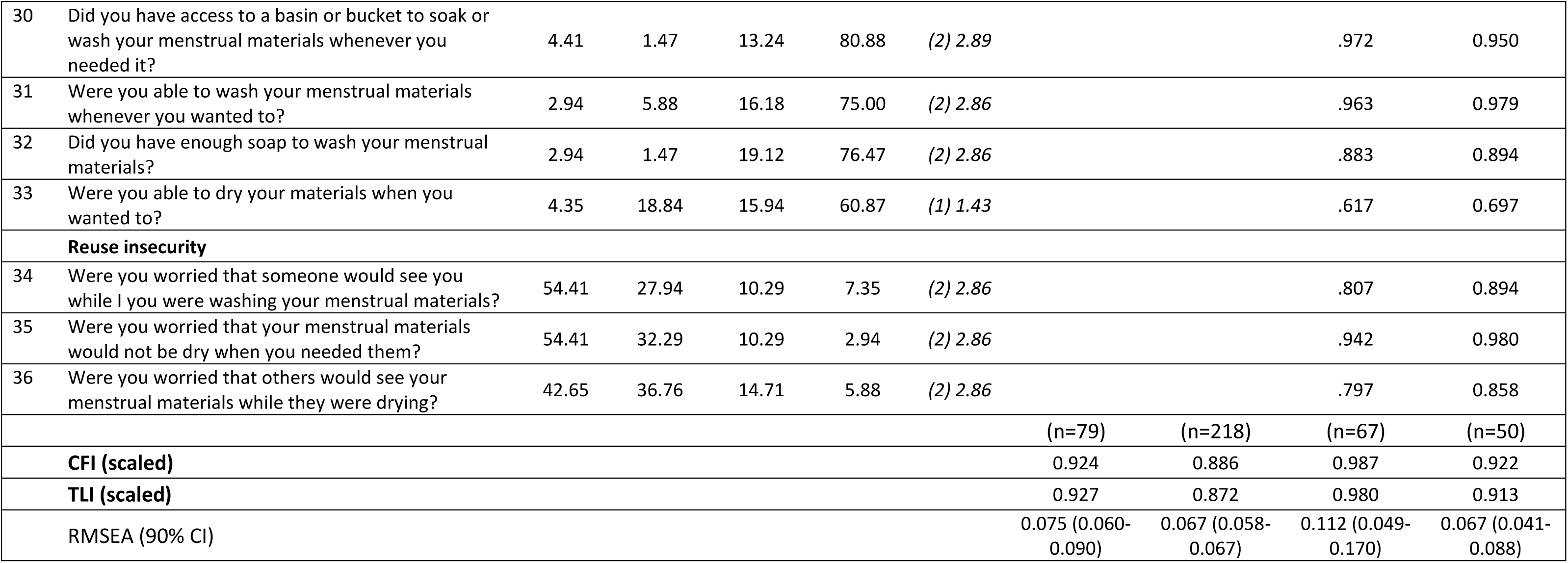
Proportion of participant responses to the full MPNS-36 questions, and confirmatory factor analysis for the original measure in *Bangla*.

### MPNS-36 internal consistency and validity in the Khulna pilot

The full original sub-scale exhibited strong internal consistency (Cronbach’s α = 0.86), with sub-scales ranging from 0.64 (for the 3-item material reliability concerns sub-scale) to 0.81 for items capturing insecurities about changing and disposal. Consistent with the cognitive interviews and further study information from the pilot sample, girls’ experiences of managing menstruation at school were rated much more poorly than experiences at home, with the transport and school environment needs sub-scale mean only 0.96, compared to a mean of 2.30 capturing girls’ satisfaction with materials and menstrual management at home (presented in Table 2 below). We found hypothesised relationships between the MPNS-36 and measures of mental health. While the MPNS total score was not associated with absenteeism during the last menstrual period, it was associated with 2.72 increased odds of not experiencing difficulties participating in class due to menstruation. The MPNS-36 was not associated with confidence managing menstruation at home, but sub-scales capturing satisfaction with materials, and with the school environment were positively associated with confidence managing menstruation at school. Findings suggest the original MPNS-36 shows good internal consistency and validity in the Khulna pilot sample.

**Table 2.**
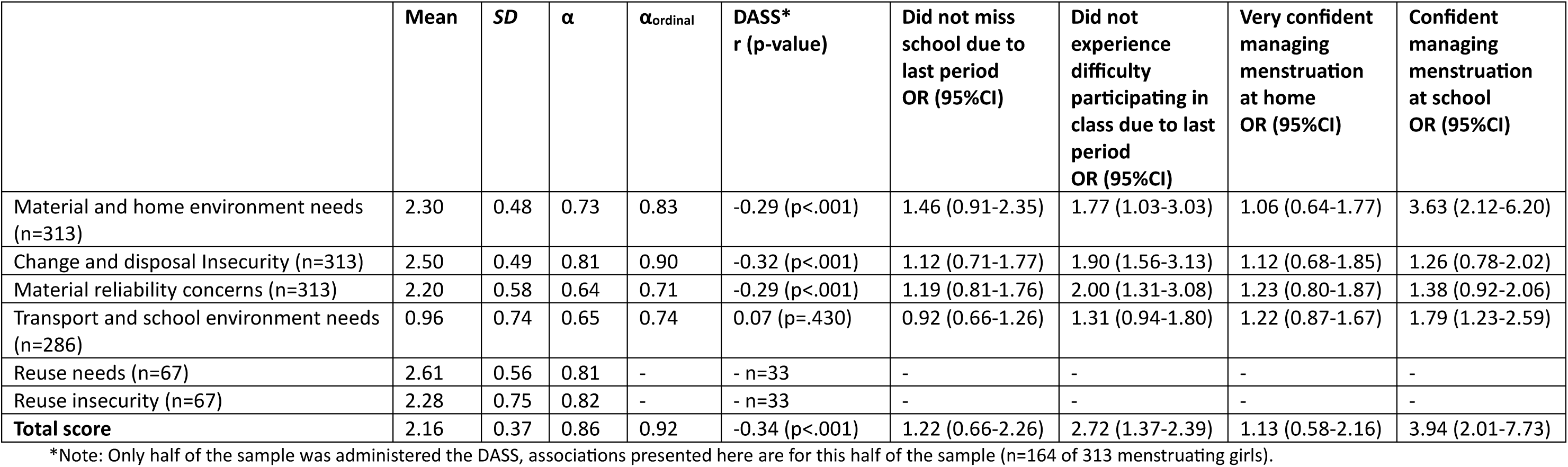
MPNS-36 total and sub-scale scores, internal consistency and associations with hypothesised correlates in the Khulna pilot data.

**Supplementary Materials 3.** Descriptive responses for each sample for hypothesised correlates

|  | Khulna Pilot, Bangladesh<br>% (n) | Khulna Cohort,<br>Bangladesh<br>% (n) | Soroti, Uganda<br>% (n) |
| --- | --- | --- | --- |
| **n** | 313 | 891 | 538 |
| **Missed school due to last menstrual period** | 44.9 (137) | 36.9 (321) | 31.1 (161) |
| **Experienced difficulty participating in class due to last menstrual period** | 47.5 (135) | 45.8 (385) | - |
| **Felt very confident managing menstruation at home** | 30.6 (96) | 26.0 (232) | Felt confident<br>80.5 (432) |
| **Felt confident managing menstruation at school** | 66.4 (207) | 48.4 (431) | 51.6 (277) |

**Supplementary Materials 3.** MPNS-SF and MPNS-R total and sub-scale scores, internal consistency and associations with hypothesised correlates across the three included surveys

| KHULNA PILOT | Mean | SD | $\alpha$ | $\alpha_{\text{ordinal}}$ | DASS<br>r (p-value)<sup>1</sup> | Did not miss<br>school due to<br>last period<br>OR (95%CI) | Did not<br>experience<br>difficulty<br>participating in<br>class due to last<br>period<br>OR (95%CI) | Very confident<br>managing<br>menstruation at<br>home<br>OR (95%CI) | Confident<br>managing<br>menstruation at<br>school<br>OR (95%CI) |
| --- | --- | --- | --- | --- | --- | --- | --- | --- | --- |
| **MPNS-36 Total** | 2.16 | 0.37 | 0.86 | 0.92 | -0.34 (p<.001) | 1.22 (0.66-2.26) | 2.72 (1.37-2.39) | 1.13 (0.58-2.16) | 3.94 (2.01-7.73) |
| **MPNS-SF Total** | 2.21 | 0.39 | 0.78 | 0.88 | -0.28 (p<.001) | 1.07 (0.60-1.90) | 2.56 (1.35-4.86) | 1.40 (0.75-2.60) | 2.91 (1.57-5.39) |
| (SF) Material and home<br>environment needs (n=313) | 2.35 | 0.54 | 0.55 | 0.67 | -0.25 (p=.001) | 1.50 (0.98-2.28) | 1.94 (1.21-3.13) | 1.49 (0.93-2.37) | 2.62 (1.66-4.13) |
| (SF) Change and disposal<br>Insecurity (n=313) | 2.56 | 0.54 | 0.82 | 0.88 | -0.33 (p<.001) | 1.24 (0.81-1.88) | 1.35 (0.87-2.10) | 1.02 (0.65-1.60) | 1.43 (0.93-2.18) |
| (SF) Material reliability concerns<br>(n=313) | 2.31 | 0.57 | 0.33 | 0.46 | -0.26 (p<.001) | 1.12 (0.76-1.67) | 1.87 (1.22-2.87) | 1.31 (0.85-2.02) | 1.56 (1.03-2.35) |
| (SF) School environment needs<br>(n=272) | 0.83 | 0.92 | 0.65 | 0.77 | 0.14 (0.086) | 0.86 (0.66-1.12) | 1.31 (1.00-1.71) | 1.26 (0.96-1.65) | 1.17 (0.87-1.56) |
| (SF) Reuse needs (n=68) | 2.68 | 0.58 | 0.81 | 0.90 | - n=33 |  |  | 0.61 (0.25-1.50) |  |
| (SF) Reuse insecurity (n=68) | 2.27 | 0.76 | 0.77 | 0.84 | - n=33 |  |  | 1.63 (0.66-4.00) |  |
| **MPNS-R Total** | 2.14 | 0.45 | 0.59 | 0.74 | -0.32 (p<.001) | 1.09 (0.66-1.83) | 2.36 (1.33-4.19) | 1.26 (0.73-2.16) | 2.32 (1.35-3.98) |

| KHULNA COHORT | Mean | SD | $\alpha$ | $\alpha_{\text{ordinal}}$ | DASS<br>r (p-value)‡ | Did not miss<br>school due to<br>last period<br>OR (95%CI) | Did not<br>experience<br>difficulty<br>participating in<br>class due to last<br>period<br>OR (95%CI) | Very confident<br>managing<br>menstruation at<br>home<br>OR (95%CI) | Confident<br>managing<br>menstruation at<br>school<br>OR (95%CI) |
| --- | --- | --- | --- | --- | --- | --- | --- | --- | --- |
| **MPNS-SF Total** | 2.08 | 0.44 | 0.78 | **0.87** | -0.36 (p<.001) | 1.88 (1.37-2.57) | 4.80 (3.36-6.86) | 2.40 (1.66-3.47) | 1.58 (1.17-2.14) |
| (SF) Material and home<br>environment needs (n=891) | 2.28 | 0.54 | 0.59 | 0.71 | -0.21 (p<.001) | 1.47 (1.14-1.89) | 2.06 (1.58-2.69) | 2.08 (1.53-2.83) | 1.09 (0.85-1.39) |
| (SF) Change and disposal<br>Insecurity (n=890) | 2.34 | 0.74 | 0.78 | 0.86 | -0.35 (p<.001) | 1.51 (1.25-1.81) | 1.70 (1.40-2.07) | 1.17 (0.95-1.44) | 1.09 (0.91-1.30) |
| (SF) Material reliability concerns<br>(n=891) | 2.01 | 0.77 | 0.43 | 0.54 | -0.25 (p<.001) | 1.37 (1.15-1.64) | 1.86 (1.54-2.24) | 1.21 (0.99-1.47) | 1.24 (1.04-1.47) |
| (SF) School environment needs<br>(n=780) | 0.86 | 0.86 | 0.51 | 0.64 | -0.08<br>(p=0.023) | 1.16 (0.97-1.38) | 1.50 (1.26-1.79) | 1.42 (1.19-1.70) | 1.66 (1.39-1.97) |
| (SF) Reuse needs (n=365) | 2.55 | 0.64 | 0.60 | 0.80 | -0.23 (p<.001) | 1.42 (1.00-2.00) | 2.27 (1.57-3.29) | 2.04 (1.24-3.37) | 1.24 (0.89-1.73) |
| (SF) Reuse insecurity (n=365) | 2.09 | 0.88 | 0.59 | 0.72 | -0.29 (p<.001) | 1.15 (0.90-1.46) | 2.19 (1.67-2.87) | 1.24-0.92-1.68) | 1.15 (0.90-1.46) |
| **MPNS-R Total** | 1.99 | 0.49 | 0.60 | 0.72 | -0.32 (p<.001) | 1.77 (1.33-2.36) | 3.65 (2.65-2.01) | 2.36 (1.68-3.30) | 1.54 (1.17-2.03) |
| **SOROTI STUDY** | **Mean** | **SD** | **$\alpha$** | **$\alpha_{\text{ordinal}}$** | **DASS**<br>**r (p-value)‡** | **Did not miss**<br>**school due to**<br>**last period**<br>**OR (95%CI)** | **--** | **Confident**<br>**managing**<br>**menstruation at**<br>**home**<br>**OR (95%CI)** | **Confident**<br>**managing**<br>**menstruation at**<br>**school**<br>**OR (95%CI)** |
| **MPNS-36 Total** <sup>2</sup> | 1.82 | 0.37 | 0.77 | 0.82 | -0.11 (p=.013) | 2.62 (1.52-4.50) | - | 4.09 (2.52-7.06) | 4.22 (2.52-7.06) |
| **MPNS-SF Total** | 1.79 | 0.40 | 0.64 | 0.72 | -0.12 (p=.005) | 2.42 (1.48-3.97) |  | 3.09 (1.75-5.45) | 4.02 (2.50-6.46) |
| (SF) Material and home<br>environment needs (n=525) | 1.97 | 0.70 | 0.60 |  | 0.06 (p=.207) | 1.11 (0.85-1.46) |  | 2.64 (1.89-3.66) | 1.74 (1.34-2.25) |
| (SF) Change and disposal<br>Insecurity (n=525) | 1.89 | 0.70 | 0.66 |  | -0.24 (p<.001) | 1.69 (1.28-2.24) |  | 0.92 (0.67-1.26) | 1.12 (0.88-1.43) |
| (SF) Material reliability concerns<br>(n=525) | 1.75 | 0.84 | 0.44 |  | -0.26 (p<.001) | 1.44 (1.15-1.81) |  | 1.10 (0.85-1.42) | 1.10 (0.90-1.35) |
| (SF) School environment needs<br>(n=525) | 1.36 | 0.91 | 0.64 |  | 0.13 (p=.003) | 1.03 (0.84-1.26) |  | 1.15 (0.91-1.46) | 1.81 (1.47-2.20) |
| (SF) Reuse needs (n=286) | 2.12 | 0.81 | 0.59 |  | -0.03 (p=.640) | 1.71 (1.25-2.34) |  | 1.88 (1.31-2.38) | 1.67 (1.23-2.27) |
| (SF) Reuse insecurity (n=285) | 1.77 | 0.80 | 0.29 |  | -0.20 (p<.001) | 1.32 (0.96-1.82) |  | 1.38 (0.95-1.99) | 0.99 (0.74-1.32) |
| **MPNS-R Total** | 1.79 | 0.47 | 0.43 | 0.48 | -0.04 (p=.310) | 1.48 (0.98-2.22) |  | 2.27 (1.41-3.65) | 3.46 (2.31-5.19) |

<sup>1</sup> Khulna Pilot: Only half of the sample was administered the DASS, associations presented here are for this half of the sample (n=164 of 313 menstruating girls) | Khulna Cohort and Soroti data. <sup>2</sup> Findings from the original study report[3], provided here for ease of comparison

